# Outcomes and Management When Using High-sensitivity Versus Conventional Cardiac Troponin in Patients With Chest Pain: A Meta-analysis

**DOI:** 10.1101/2023.10.27.23297701

**Authors:** John Christopher A. Pilapil, Paula Victoria Catherine Y. Cheng, Paul Anthony O. Alad, Jaime Alfonso M. Aherrera

## Abstract

**Background:** Cardiac troponin (cTn) measurement is pivotal in diagnosing and managing chest pain. High-sensitivity cTn (hs-cTn) assays are slowly supplanting conventional cardiac troponin (c-cTn) use, allowing earlier identification of high-risk patients and diagnosis of myocardial infarction (MI). This improved sensitivity is said to come at the cost of reduced specificity for MI. However, there is a paucity of data regarding the exact impact of this change in patient management and outcomes.

**Objectives:** To compare outcomes and management of patients with chest pain when using hs-cTn versus c-cTn.

**Methods:** We conducted a systematic search of studies in all languages across various databases. Inclusion criteria for studies were (1) observational or randomized trials, (2) included adult patients presenting with chest pain, (3) use of hs-cTn or c-cTn in diagnosis, (3) reported data on any of the pre-defined outcomes. The primary outcome was all-cause mortality and secondary outcomes were major adverse cardiovascular events (MACE), MI following index admission, coronary angiography, and revascularization. Study quality was appraised using the Cochrane Tool for Assessing Risk of Bias for randomized trials and the Newcastle-Ottawa Quality Assessment scale for observational studies. Outcomes were analyzed using Review Manager (RevMan) 5.3, employing Mantel-Haenszel analysis of random effects to compute for relative risk and odds ratio.

**Results:** Pooled analysis from 5 studies showed that among patients with chest pain, those for whom hs-cTn over c-cTn was used had no difference in all-cause mortality (RR 1.01, 95% CI 0.92-1.12, p=0.82, I^2^=0%), a significant decrease in MI (RR 0.74, 95% CI 0.63-0.87, p=0.0003) and a trend towards increase in MACE (RR 1.08, 95% CI 1.00-1.16, p=0.04, I^2^=0%). They were more likely to undergo coronary angiogram (OR 1.52, 95% CI 1.02-2.28, p=0.04) and revascularization (OR 1.34, 95% CI 1.03-1.75, p=0.03).

**Conclusion:** Use of hs-cTn over conventional cTn led to higher rates of coronary angiography and revascularization. While there was significant reduction in myocardial infarction, there was no reduction in all-cause mortality and even a trends towards increased MACE.

## I. INTRODUCTION

### Acute Chest Pain

Acute chest pain remains to be one of the most common complaints for seeking care in the emergency department (ED). Next to injuries, chest pain is the second most common complaint in the United States – accounting for over 7 million or 4.7% of all annual ED visits and almost 10% of approximately 100 million non-traumatic visits^1,2^. The lifetime prevalence of chest pain is estimated at 20-40%, with more women than men experiencing the symptom. More than 60% of patients at the ED with this presentation are admitted for further evaluation^3,4^.

Pathologies involving the heart, vasculature, lungs, pleura, mediastinal structures, gastrointestinal tract, musculoskeletal system, and nervous system may all present with chest discomfort^5^. Although the determined cause of patients at the ED with acute chest pain is typically non-cardiac, ischemic heart disease (IHD) still represents 31% of all etiologies and is the most common serious, life-threatening cause^6^. Chest pain, in turn, remains to be the most common symptom of the entire spectrum of coronary artery disease (CAD) – both acute and chronic disease – in men and women^7^. Of all patients presenting to the ED with chest pain, an estimated 5-15% will have acute coronary syndrome (ACS).

Thus, chest pain continues to prove itself as a diagnostic challenge, requiring thorough clinical evaluation and prudent, prompt use of diagnostics in both the outpatient and ED setting. Differentiating patients with ACS and CAD or other life-threatening conditions – cardiac and non-cardiac alike – from those with non-cardiovascular, non-life-threatening chest pain is of paramount importance to the attending clinician.

### Cardiac Troponin Assays: High Sensitivity vs. Conventional

Patients presenting with acute chest pain suspected to have ACS necessitate measurement of biomarkers for myocardial injury^8,9,10,11^. Cardiac troponin (cTn: T or I; cTnT or cTnI) is the preferred biomarker. cTnI and cTnT are cardiac regulatory proteins involved in calcium-mediated actin and myosin interaction^12^. Since they are products of genes expressed almost exclusively by cardiac tissue, cTn’s are potentially specific cardiac isoforms and essentially unique to the heart^13,14^. In their clinical use, both cTnT and cTnI have shown equal utility and diagnostic capability^15^. The once popular creatinine kinase MB isoenzyme (CK-MB) is no longer recommended due to its lack of sensitivity and specificity^8–11,12–14^.

Assays for cTn measurement are done using enzyme-linked immunosorbent testing techniques and are available with varying sensitivities to levels of the biomarker of interest [16]. The limit of detection is defined as the lowest, detectable cTn concentration that is accurately measured in a sample containing low cTn concentration^17–18^. Compared with the older conventional cTn (c-cTn) assays, high-sensitivity cTn (hs-cTn) assays have the distinction of being able to detect cTn values above the limit of detection in at least 50% of normal individuals^19^.

The advent of modern hs-cTn assays enable earlier detection of MI, essentially shifting the conventions for workup of chest pain by allowing early “rule-out” strategies^20–23^. Due to their greater precision at the lower range of measurable troponin level, hs-cTn as opposed to c-cTn assays allow: (1) determination if the cTn in a blood sample is elevated or above the upper limit of normal, and moreover, (2) reliable detection of absolute changes in samples obtained serially at presentation and hours later^21–27^. The more sensitive examination by hs-cTn assays not only of absolute cTn levels, but dynamic changes as well has thus shortened the time interval to the next measurement from a historical practice of 6-12 or more hours to 1-2 hours. All of this while still achieving good negative predictive values of ≥ 99%^8–9,24–27^. Such changes in clinical algorithms and strategies have likewise facilitated discharging patients on the basis of a single cTn value at presentation with reasonable safety^9,11,28^.

The preferential use of hs-cTn assays for biomarker measurement has likewise allowed for the detection of potentially fatal and intervenable disease in a broader range of patients. The heightened sensitivity for detection of ACS, however, also includes patients with other causes of cTn elevation^28–30^. Reasons of elevation of cTn values due to myocardial injury appear to be all-encompassing and may include injury due to myocardial ischemia by virtue of CAD and atherosclerotic plaque disruption with thrombosis, myocardial ischemia due to oxygen supply/demand imbalance, and other “miscellaneous” causes of myocardial injury – such as non-CAD cardiac conditions and various systemic diseases^20^. A previous collaborative meta-analysis has suggested that hs-cTn assays improve early sensitivity and negative predictive value for the diagnosis of ACS and acute MI, but at the expense of specificity and positive predictive value^30^.

### Importance and Significance of the Study

Hs-cTn assays are now preferred over c-cTn assays, as recommended by the Fourth Universal Definition of Myocardial Infarction. Here, myocardial injury is defined as the detection of an elevated cTn value above the 99^th^ percentile upper reference limit. Furthermore, the presence of a rise and/or fall of cTn values is said to be indicative that the injury is acute^20^. The recommendation for using hs-cTn over c-cTn assays in consistent in practically all guidelines detailing the diagnosis and management of myocardial infarction (MI)^8–11,20^. With this, the use of hs-cTn continues to replace c-cTn in the approach to patients presenting with acute chest pain at the ED.

As discussed, the advent of hs-cTn assays has allowed earlier identification of high-risk patients and diagnosis of ACS among the general population presenting with chest pain. This improved sensitivity, however, seemingly comes at the cost of a reduced specificity for MI. While hs-cTn has slowly supplanted c-cTn in practice, there is a paucity of data as to the exact impact of this change in patient management and outcomes. Prior data suggests that hs-cTn use over c-cTn may improve prognosis, but data is wanting to definitively confirm this^31^.

The necessity of looking into the particularities of hs-cTn vs. c-cTn use in patients with chest pain is perhaps made all the important by the fact that in recent past, numerous advances and trials have yielded paradigm shifts in the way we approach patients with chest pain and subsequently manage those confirmed to have ACS – whether via conservative/medical or invasive strategies. Our study is the first of its kind to consolidate data on the management and outcomes of hs-cTn vs. c-cTn use in adult patients presenting at the ED with chest pain.

## II. METHODS

### Literature Search

We conducted a systematic search of studies until August 1, 2023 that compared outcomes and management of patients with chest pain using hs-cTn versus conventional cTn. Meta-analyses, systematic reviews, randomized controlled trials, cohort studies, and observational studies on the following electronic databases were eligible for inclusion: PubMed, Cochrane Library, MEDLINE, EMBASE, WHO Network of Collaborating Clinical Trial Registers, ScienceDirect, and ClinicalTrials.gov from US National Institutes of Health. The following free text and MeSH terms were used in our search strategy: *high sensitivity cardiac troponin, conventional cardiac troponin, chest pain, chest discomfort, angina, acute coronary syndrome,* and *myocardial infarction.* Search for available literature PubMed was further refined by using high-sensitivity method filters to sift out applicable study designs. Retrieved articles’ reference lists were likewise examined to identify additional potential studies for inclusion. We attempted to contact study authors and known experts in the field to search for additional published and unpublished data alike.

### Study Inclusion

Two authors independently assessed the retrieved articles to determine those for inclusion in this meta-analysis. We determined the following pre-specified inclusion criteria for studies:

- Population: Adult patients ≥ 19 years old presenting to the emergency department with chest pain or discomfort
- Interventions: Use of hs-cTn versus conventional cTn
- Outcomes:
- `o` Primary efficacy outcome: All-cause mortality
- `o` Secondary efficacy outcomes: Major adverse cardiac events, myocardial infarction after index admission
- `o` Management strategies: Rate of performance of coronary angiography, rate of revascularization
- Study Type: Randomized controlled trials, prospective and retrospective cohort and observational studies

## Data Collection and Analysis

### Assessment of Risk of Bias in Included Studies

Two reviewers independently assessed the validity and risk of bias of retrieved articles. The Newcastle-Ottawa Quality Assessment Form was utilized for cohort studies and case control studies^32^. Articles were determined to be of good, fair, or poor quality. Risk of bias and study quality tables were constructed. Disputes and disagreements with regard to the article appraisal was settled by discussion and consensus of the reviewers with a third author who served as arbiter.

### Data Extraction and Management

Two reviewers independently extracted the data using a uniform data extraction form. Each eligible study was reviewed by both authors and any disputes and disagreements were addressed through discussion and arbitration with a third reviewer. The following data were extracted from each study: all-cause mortality, major adverse cardiovascular events, myocardial infarction following index admission, performance of coronary angiography, and revascularization.

### Measures of Treatment Effect

Data was analyzed using Review Manager 5, utilizing the Mantel-Haenszel test statistical method and random effects model. Clinical outcomes were calculated for their relative risks, while management strategies employed were calculated for their odds ratios. Data from each trial included was synthesized and combined for cases deemed appropriate. A 95% confidence interval was also presented with each result.

### Assessment of Heterogeneity

Heterogeneity of studies was assessed by analyzing the study characteristics, and similarities or differences between participants, interventions, results, and methodologies. Heterogeneity was determined using the I^2^ and chi-square measures of heterogeneity, setting a value of more than or equal to 50% I^2^ and less than or equal to 10% p-value to indicate statistically significant heterogeneity.

### Data Analysis and Synthesis

Data analysis and synthesis was done through the Review Manager 5 software. Relative risks, risk ratios, and odds ratios of dichotomous variables were calculated using the random effects model.

## III. RESULTS

### INCLUDED AND EXCLUDED STUDIES

Initial search and identification of studies yielded 62 non-duplicate records from the various databases and registries listed in this study’s methods section, which were then screened by the reviewers using the set selection criteria for studies. A total of 52 articles were excluded after screening the studies’ titles and abstracts. 10 articles were retrieved for full-text review. Among these, 6 were further excluded due to non-compatibility with this study’s selection criteria – three of the articles did not include any of our desired and pre-specified outcomes, two articles had different investigated interventions, and one article analyzed a different population compared to our research objectives. Among the remaining 4 articles, another 1 was identified via citation searching and deemed eligible. A total of 5 studies were included in the final list of studies for the meta-analysis. The PRISMA workflow chart^33^ showing article and study identification, screening, inclusion, and exclusion may be found in *Supplemental Appendix A*.

### STUDY CHARACTERISTICS AND QUALITY

Five retrieved studies (n = 504,123) met our inclusion criteria and were included in the final analysis^34–38^. All of the studies were observational in design. Two of the retrieved articles utilized a prospective design, while the other three were retrospective in nature. Individual study populations ranged from 3,492 to 394,910. No randomized trials were found by our search for articles. The populations involved for four of the studies were adult patients presenting to the emergency department with chest pain and suspected ACS. One study looked at adult presents presenting with chest pain who were subsequently diagnosed for the first time with ACS^36^. All of the included studies reported at least one of our specified outcomes of interest. Follow-up of patients ranged from as short as 30 days to as long as nearly 3 years. All of the studies were of good quality by the Newcastle-Ottawa Scale. Table 1 summarizes the characteristics of included studies. The details of risk of bias assessment may be found in *Supplemental Appendix B*.

**Table 1.** Characteristics of Included Studies.

| **Author** | **Study design** | **Location** | **N** | **Population** | **Intervention Group** | **Control** | **Pertinent Outcomes** | **Duration / Follow-up** |
| --- | --- | --- | --- | --- | --- | --- | --- | --- |
| Kaambwa et al. (2017) [34] | Prospective, multicenter, pragmatic | Adelaide, South Australia | 1937 | Patients with low and intermediate risk chest pain presenting to emergency services for evaluation of suspected ACS | hs-cTn | c-cTn | Adverse clinical outcomes (all-cause mortality, recurrent ACS beyond the first 24 hours up to 12 months)<br>Quality of life<br>Economic cost analysis | Follow-up: 12 months |
| Lau et al. (2018) [35] | Retrospective, observational | Ontario, Canada | 394910 | Patients aged 40-105 who presented to the emergency department with chest pain and received an ECG | hs-cTn | c-cTn | All-cause mortality<br>Myocardial infarction<br>Angina<br>Death, MI, or angina | Follow-up 30, 90, and 365 days post-discharge |
| Odqvist et al. (2018) [36] | Retrospective cohort | Sweden | 87879 | Patients hospitalized for a first MI during the period from 2009-2013 in Sweden and were diagnosed using either hs-cTn or c-cTn at hospitals where the 99<sup>th</sup> percentile value was used as the | hs-cTn | c-cTn | All-cause mortality<br>Reinfarction<br>Coronary angiography<br>Revascularization | Duration: 2009-2013 |

|  |  |  |  | decision limit for MI |  |  |  |  |
| --- | --- | --- | --- | --- | --- | --- | --- | --- |
| Sanchis et al. (2014) [37] | Prospective cohort | Valencia, Spain | 1372 | Patients who presented at the emergency department with acute chest pain | hs-cTn | c-cTn | MACE (All-cause death, readmission for MI, readmission for UA, and post-discharge revascularization)<br>All-cause death<br>MI<br>UA readmission<br>Post-discharge revascularization<br>Coronary angiogram<br>PCI<br>CABG<br>Revascularization | Duration: c-cTn from Mar. 1, 2008 to July 1, 2010)<br>hs-cTn from Nov. 1, 2010 to Mar. 1, 2013<br>Follow-up: 6 months |
| **Zachoval (2020)** [38] | Retrospective cohort | Germany | 18025 | Patients with chest pain and/or suspected ACS where at least one troponin measurement was obtained | hs-cTn T | c-cTn I | Mortality<br>Acute MI<br>Coronary angiography<br>PCI | Duration: c-cTn I from October 2016 to April 2017, hs-cTn T from October 2017 to April 2018 |

*ACS – acute coronary syndrome, CABG – coronary artery bypass grafting, ECG – electrocardiogram, MACE – major adverse cardiac events, MI – myocardial infarction, PCI – percutaneous coronary intervention, UA – unstable angina*

## OUTCOMES

### Primary Outcome

Four studies (n = 502,116) were included in the initial analysis (Fig. 1). Pooled data showed that the use of hs-cTn over c-cTn had no effect on all-cause mortality (RR 0.94, 95% CI 0.76-1.16, p = 0.54). Results, however, were with significant heterogeneity (I^2^ = 90%). The study authors identified the study by Odqvist et al^36^ to be a contributor to the noted heterogeneity. Having included patients already diagnosed with myocardial infarction renders their study population already of high risk at the onset. The effect of using hs-cTn that would lead to earlier and perhaps more aggressive interventions, therefore, was postulated to contribute to heterogeneity by skewing benefit towards the use of hs-cTn.

**Figure 1.**
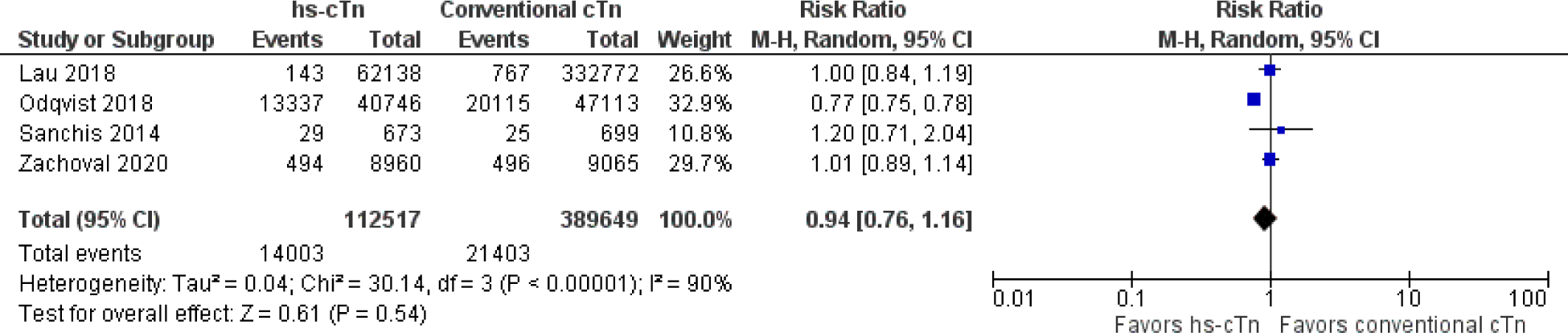
Forest Plot reporting pooled risk ratio with 95% confidence interval for all-cause mortality, comparing patients with chest pain with hs-cTn vs. conventional cTn used as part of diagnostic strategy.

An analysis adjusting for the identified source of heterogeneity (Fig. 2) was thus carried out, identifying the study of Odqvist et al due to its population being of higher risk for cardiovascular events compared to the others. From this, three studies (n = 414,307) were included in the final analysis. This revealed that, indeed, the use of hs-cTn over c-cTn did not lead to differences in all-cause mortality (RR 1.01, 95% CI 0.92-1.12, p=0.82).

**Figure 2.**
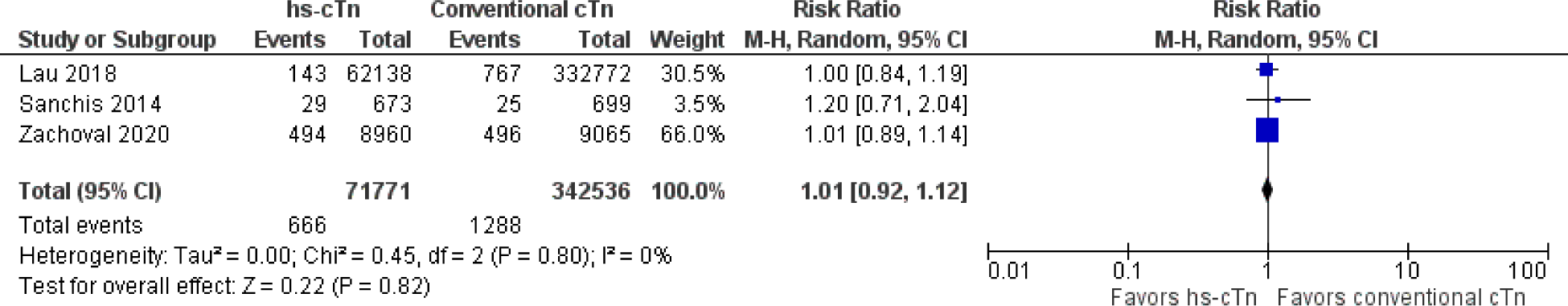
Forest Plot reporting pooled ratio with 95% confidence interval for all-cause mortality, after adjusting for heterogeneity by excluding the study by Odqvist et al (2018).

### Secondary Outcomes

#### Secondary Clinical Outcomes

##### Major Adverse Cardiovascular Events (MACE)

Two studies (n = 3,309) reporting on MACE were included in the analysis (Fig. 3). Results showed that the use of hs-cTn as opposed to c-cTn did not lead to a significant reduction in MACE, without significant heterogeneity (RR 1.08, 95% CI 1.00-1.16, p=0.04, I^2^=0%). They even noted a signal towards harm that would appear to favor using c-cTn.

**Figure 3.**
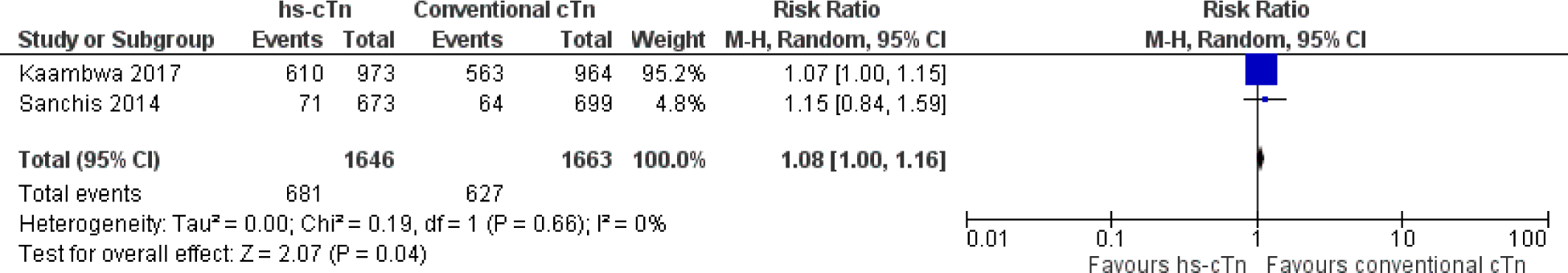
Forest Plot reporting pooled risk ratio with 95% confidence interval for major adverse cardiovascular events, comparing patients with chest pain with hs-cTn vs. conventional cTn used as part of diagnostic strategy.

##### Myocardial Infarction on Follow-up

A total of 4 studies (n = 492,942) were included in this analysis (Fig. 4), which showed that there was an overall statistically significant decrease in the incidence of myocardial infarction on subsequent follow-ups of patients for whom hs-cTn was used instead of c-cTn as part of the diagnostic strategy (RR 0.74, 95% CI 0.63-0.87, p=0.0003), but with significant heterogeneity of these results (I^2^=87%).

**Figure 4.**
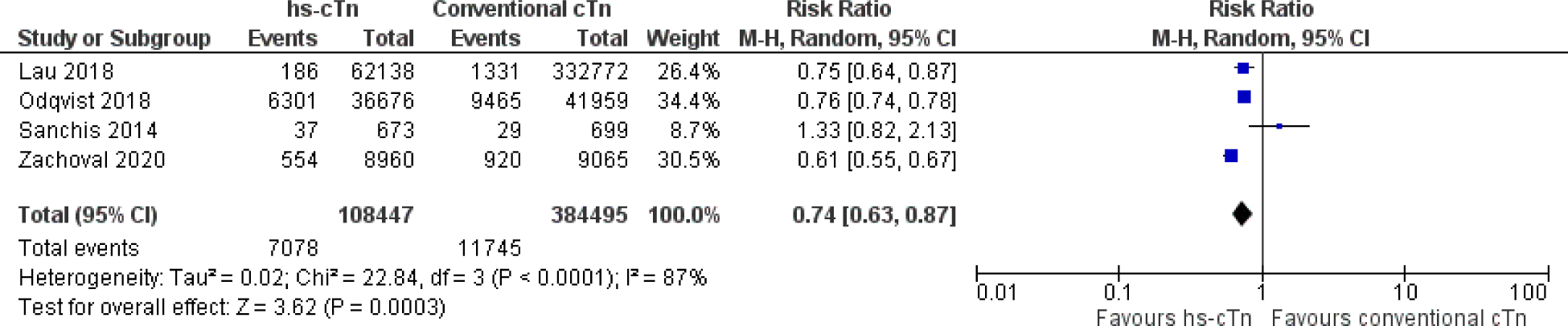
Forest Plot reporting pooled risk ratio with 95% confidence interval for myocardial infarction on follow-up, comparing patients with chest pain with hs-cTn vs. conventional cTn used as part of diagnostic strategy.

#### Management of Patients

##### Performance of Coronary Angiogram

With three studies (n = 107,276) included in the final analysis (Fig. 5), results reveal that there was a statistically significant increase in the performance of coronary angiogram among patients for whom hs-cTn was used over c-cTn (OR 1.52, 95% CI 1.02-2.28, p=0.04). Results were with heterogeneity (I^2^=98%).

**Figure 5.**
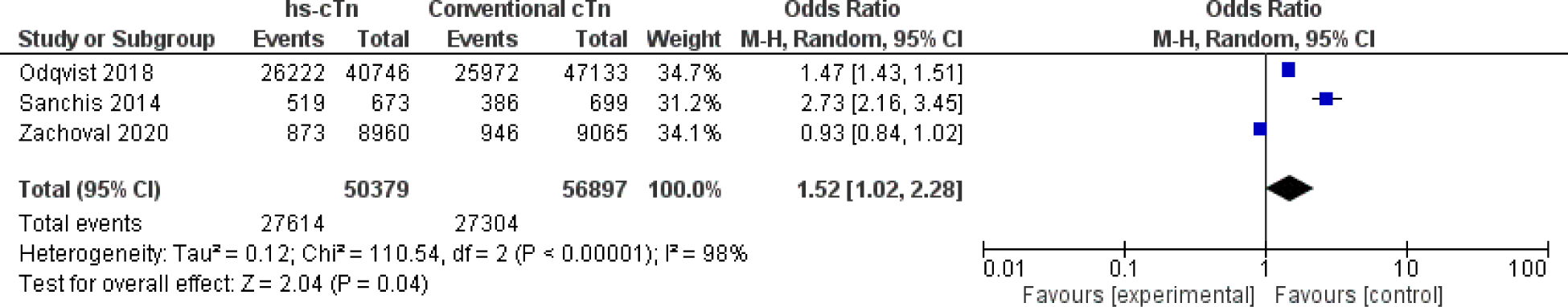
Forest Plot reporting pooled odds ratio with 95% confidence interval for performance of coronary angiography, comparing patients with chest pain with hs-cTn vs. conventional cTn used as part of diagnostic strategy.

##### Performance of Revascularization

Three studies (n = 107,276) were included in the analysis (Fig. 6). There was a statistically significant increase in revascularization among patients for whom hs-cTn was used instead of c-cTn (OR 1.34, 95% CI 1.03-1.75, p=0.03), but with heterogeneous results (I^2^=93%).

**Figure 6.**
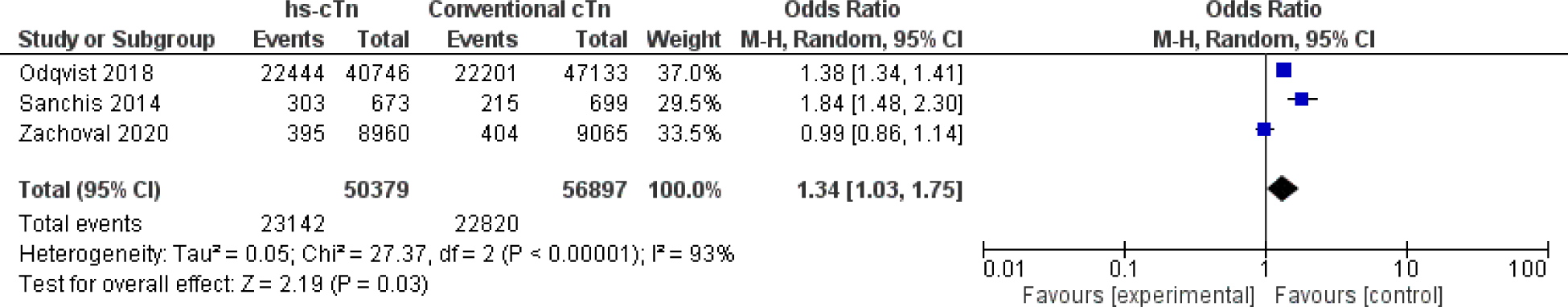
Forest Plot reporting pooled odds ratio with 95% confidence interval for revascularization, comparing patients with chest pain with hs-cTn vs. conventional cTn used as part of diagnostic strategy.

## IV. DISCUSSION

Our meta-analysis, which included 5 cohort studies involving 504,123 patients, is the first to consolidate and analyze data pertaining to the management and outcomes of patients presenting to the ED with chest pain when using hs-cTn vs. c-cTn as part of the diagnostic strategy and patient approach. Our analysis shows the following results that may be highlighted into two points. First, the use of hs-cTn when compared with c-cTn in these patients led to more interventions being performed, with higher rates of coronary angiography and revascularization. Secondly, with this increased performance of diagnostic and therapeutic procedures, there was a significant decrease in MI on follow-up, but no reduction in all-cause mortality and even a signal towards increased major cardiac events.

### Effects on Patient Management

This study shows that using hs-cTn over c-cTn assays led to statistically significant increased odds for the performance of coronary angiogram (OR 1.52, 95% CI 1.02-2.28, p=0.04). There was likewise an increase in the odds of revascularization (OR 1.34, 95% CI 1.03-1.75, p=0.03) – whether via coronary artery bypass grafting (CABG) or percutaneous coronary intervention (PCI), inclusive of balloon angioplasty with or without stent implantation.

A possible explanation to this observed phenomenon is that an early invasive approach (via coronary angiogram with the intent to revascularize diseased vessels) is recommended for patients with acute coronary syndrome diagnosed to have myocardial infarction and also in those with a significant rise and/or fall in cTn values on serial measurement^8–11^. As we have detailed in our review of literature, hs-cTn unveils cTn elevation in some patients who otherwise would initially pass undetected using a conventional assay^18,19,29,30^. Indeed, in an early study analyzing the effects of shift from c-cTn to hs-cTn use in patients at the ED with chest pain, there was a noted increase in prevalence of MI from 18% to 22%^39^. Initial concern was hence expressed that the introduction of hs-cTn assays would be followed by a concomitant increase in MI’s [36]. Subsequent studies, however, have had conflicting and ambivalent results. While some did not any change in reported MI’s with the use of hs-cTn assays over c-cTn assays^40^, others demonstrated higher MI prevalence^36,37,41^ and in some studies, even a decrease in the number of MI’s reported^42^. It is important to note that because the kinetics of troponin rise and fall are multifactorial, there is still no definitive criteria for what cTn change or “delta” is significant^43^. Since part of the diagnosis of MI is established by the pattern of rise and fall in cTn and not just cTn elevation alone, this may have contributed to the differences in the above observations with regard to the effect of hs-cTn use on MI prevalence.

Apart from the diagnosis of MI and demonstrable rise and/or fall in cTns, early performance (within 24 hours of diagnosis) of coronary angiogram is also recommended patients deemed high risk^8–11^ via risk calculators such as the GRACE score for ACS^44^ and TIMI score for non-ST elevation MI^45^. As these risk calculators frequently include elevated cTn’s as predictors that elevate the risk for events, it is possible that the more sensitive detection of even low levels of cTn by high sensitivity assays lead to higher risk scores for ACS patients overall, although there have yet to be any studies looking into this.

Significant heterogeneity was seen in our pooled analysis of results for both coronary angiogram and revascularization performance. Potential sources of this may be the wide range in time period in which the included studies were performed. These studies were likewise done across different countries. It may be possible that the recommendations and practices for ACS significantly differed across different eras, locales, and per-hospital or institution predicaments – thus leading to heterogeneity in results for both patient management odds and outcome risks.

### Effects on Patient Outcomes

Our results show that among patients with chest pain for whom hs-cTn over c-cTn assay was used, there was no difference in all-cause mortality after adjusting for sources of heterogeneity (RR 1.01, 95% CI 0.92-1.12, p=0.82, I^2^=0%). Meanwhile, a significant decrease in MI was observed (RR 0.74, 95% CI 0.63-0.87, p=0.0003) and quite strikingly, there was a trend towards increase in MACE (RR 1.08, 95% CI 1.00-1.16, p=0.04, I^2^=0%).

Our results showing a decrease in risk of MI on follow-up are seemingly consistent with available evidence that, particularly among symptomatic CAD patients, revascularization is beneficial in relieving symptoms and potentially reducing the risk of future MI^46–47^. The noted signal for harm and higher MACE could potentially be correlated with our findings of increased performance of coronary angiography and revascularization. PCI and CABG have concomitant risk for major complications – such as death, MI, or stroke – and minor complications – such as transient ischemic attacks, vascular complications, contrast-induced nephropathy, and angiographic complications^48–50^. In theory, higher performance of procedures would yield higher prevalence of its possible complications. This might have been the case we see for our results on MACE.

Ultimately, there was no reduction in all-cause mortality when using hs-cTn over c-cTn. In light of increased performance of angiography and revascularization and reduced MI but a trend towards increased MACE, it is important to emphasize the necessity to weigh and carefully analyze the implications of preference for hs-cTn assay use in approaching with chest pain. Benefits, risks, and costs must be taken into consideration. A prior study in Australia already demonstrated that hs-cTn use had a tendency for higher health costs. In spite of fewer adverse clinical outcomes, the cost effectiveness of hs-cTn over c-cTn use was high at $108,552 per adverse clinical outcome avoided^34^. Increase in resource use leading to increased costs and higher exposure of patients to potential complications – even with the possibility for certain benefits – is a concern that must not go unnoticed. It is our recommendation that more carefully and well-designed studies should be carried out, looking into the implications on patient management, resource use, and outcomes of using hs-cTn over c-cTn.

### Study Limitations

This investigation included only articles published in English. Furthermore, there were no randomized trials found and all studies included were only observational in design. Some of the included articles were followed-up patients who were managed before more recent changes in diagnostic and therapeutic interventions for acute MI as well.

## V. CONCLUSION

Among patients presenting with chest pain, the use of hs-cTn when compared with c-cTn led to higher rates of performing coronary angiography and revascularization. While there was a significant decrease in MI on follow-up, there was no reduction in all-cause mortality and even a signal towards increased MACE.

## CLINICAL PERSPECTIVES

### CLINICAL COMPETENCIES

- Medical Knowledge: Our study is the first of its kind to consolidate data on the management and outcomes of hs-cTn vs. c-cTn use in adult patients presenting at the ED with chest pain.

### TRANSLATIONAL OUTLOOK

- Competency in Medical Knowledge: The use of hs-cTn vs. c-cTn in patients with acute chest pain allows for earlier diagnosis of acute MI, risk stratification of high risk patients. The higher sensitivity of this testing strategy is said to be at the expense of reduced specificity.
- Translational Outlook 1: The results of this meta-analysis show that using hs-cTn over c-cTn in diagnosing patients with acute chest pain leads to more aggressive management.
- Translational Outlook 2: With more aggressive management, there is a reduction in myocardial infarction on follow-up, but no effect on all-cause mortality and no significant reduction in MACE was noted.
- Translational Outlook 3: The cost-efficiency in using hs-cTn over c-cTn to work up patients with acute chest pain. The outcomes we are able to achieve may be at the expense of higher cost due to more aggressive management of patients.

## Data Availability

Data will be made available upon the request of anyone who wishes to access it.

## ABBREVIATIONS

IHD: Ischemic heart disease
CAD: Coronary artery disease
ACS: Acute coronary syndrome
MI: Myocardial infarction
cTn: T or I; cTnT or cTnI: Cardiac troponin
c-cTn: Conventional cTn
hs-cTn: High-sensitivity cTn

## DECLARATIONS

### Availability of data and materials

Data from the articles primarily used in this meta-analysis may be readily accessed through PubMed.

### Authors’ contributions

- JP is the primary author and carried out data acquisition, analaysis, interpretation of the data, and discussion writeup.
- PC contributed to data acquisition, article appraisal using the Newcastle Ottawa Scale, and data analysis. She likewise wrote part of the discussion and review of literature.
- PA contributed to data acquisition and analysis. He contributed to substantially proofreading the manuscript as well.
- JA came up with the research idea and served as the arbiter for any disagreements in article appraisal of the other authors. He likewise contributed substantially to the interpretation of data.

## SUPPLEMENTAL MATERIALS

**Appendix A.**
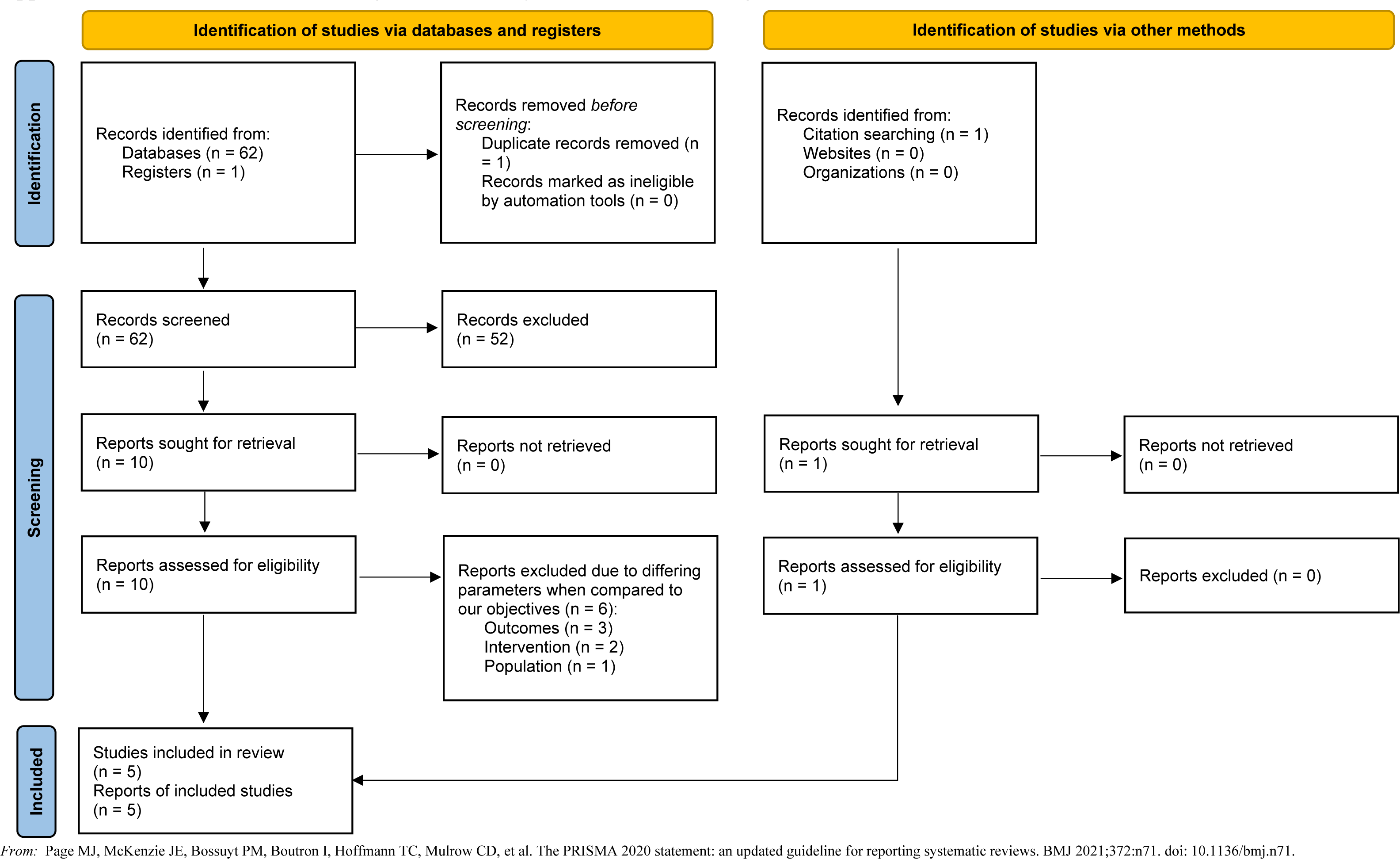
PRISMA flow chart showing article and study identification, screening, inclusion, and exclusion.

**Appendix B.** Assessment of risk of bias using the Newcastle-Ottawa Scale (NOS) for assessing the quality of nonrandomized studies in meta-analyses.

| Study | Selection |  |  |  | Comparability | Outcome |  |  | Quality |
| --- | --- | --- | --- | --- | --- | --- | --- | --- | --- |
|  | Representativeness of the exposed cohort | Selection of the non-exposed cohort | Ascertainment of exposure | Outcome of interest not present at start of study | Comparability of cohorts on the basis of the design or analysis | Assessment of outcome | Adequacy of follow-up duration | Adequacy of completeness of follow-up |  |
| **Kaambwa et al. (2017)<sup>34</sup>** | ☐ | ☐ | ☐ | ☐ |  | ☐ | ☐ | ☐ | **7** |
| **Lau et al. (2018)<sup>35</sup>** | ☐ | ☐ | ☐ | ☐ |  | ☐ | ☐ | ☐ | **7** |
| **Odqvist et al. (2018)<sup>36</sup>** |  | ☐ | ☐ | ☐ | ☐ ☐ | ☐ | ☐ | ☐ | **8** |
| **Sanchis et al. (2014)<sup>37</sup>** | ☐ | ☐ | ☐ | ☐ | ☐ ☐ | ☐ | ☐ | ☐ | **9** |
| **Zachoval (2020)<sup>38</sup>** | ☐ | ☐ | ☐ | ☐ | ☐ | ☐ | ☐ | ☐ | **8** |

